# A cell phone data driven time use analysis of the COVID-19 epidemic

**DOI:** 10.1101/2020.04.20.20073098

**Authors:** Eli P. Fenichel, Kevin Berry, Jude Bayham, Gregg Gonsalves

## Abstract

Transmission of the SAR-COV-2 virus that causes COVID-19 is largely driven by human behavior and person-to-person contact. By staying home, people reduce the probability of contacting an infectious individual, becoming infected, and passing on the virus. One of the most promising sources of data on time use is smartphone location data. We develop a time use driven proportional mixing SEIR model that naturally incorporates time spent at home measured using smartphone location data and allows people of different health statuses to behave differently. We simulate epidemics in almost every county in the United States. The model suggests that Americans’ behavioral shifts have reduced cases in 55%-86% of counties and for 71%-91% of the population, depending on modeling assumptions. Resuming pre-epidemic behavior would lead to a rapid rise in cases in most counties. Spatial patterns of bending and flattening the curve are robust to modeling assumptions. Depending on epidemic history, county demographics, and behavior within a county, returning those with acquired immunity (assuming it exists) to regular schedules generally helps reduce cumulative COVID-19 cases. The model robustly identifies which counties would experience the greatest share of case reduction relative to continued distancing behavior. The model occasionally mischaracterizes epidemic patterns in counties tightly connected to larger counties that are experiencing large epidemics. Understanding these patterns is critical for prioritizing testing resources and back-to-work planning for the United States.

## Introduction

Physical (social) distancing in the form of “staying home” is a critical tool to prevent COVID-19 mortality, and will remain so until pharmaceutical therapies or vaccines are widely available. The modeling consensus is that some version of distancing will need to persist for months, not weeks (Anderson et al., 2020; Ferguson et al., 2020). However, strong distancing can ultimately give way to isolating infectious individuals and targeted distancing measures with sufficient contact tracing capacity (Kissler et al., 2020). A new tool in the epidemiological modeling of distancing is time use data from smartphone devices. Time use provides a measuring contact risk with a recognizable unit that has metric properties unlike arbitrary distancing indexes. Here we build on prior epidemiological time use modeling (Bayham and Fenichel, 2016; Bayham et al., 2015; Berry et al., 2018) to adapt the common SEIR framework to a dynamic time use structure that enables differential behavior by health status in order to incorporate smartphone tracking data into a model of the COVID-19 epidemic for every county in the United States. We use these results to characterize the variation in the epidemic and suggest prioritization for testing and targeting for a “back to work” strategy across the United States.

The standard SEIR compartmental epidemiological model does not have a structure for incorporating dynamic behavior that responds to the state of the epidemic (Fenichel et al., 2011). However, behavior is changing rapidly, and there appear to be strong feedbacks between the COVID-19 epidemic and people’s behavior (Malik et al., 2020; Villas-Boas et al., 2020). In prior research, we developed an economic-epidemiological model based on a time-varying conditional proportional mixing structure (Fenichel, 2013; Fenichel et al., 2011) that enables physical distancing behavior to vary based on health state and respond to the state of the epidemic. This structure can readily be adapted to time use with heterogeneous mixing models using empirical time allocation-based contact structures that can be updated within a simulation (Bayham et al., 2015). These structures can be calibrated using time allocation of Americans as measured by their smartphones’ locations.

The model, which treats all counties independently, suggests substantial heterogeneity in the COVID-19 epidemic across the United States. The model suggests that some counties are likely still experiencing near exponential growth while in other counties cases have likely peaked. In some cases the peak is due to the county’s demographic structure. In others it is due to behavior. In case where the peak is causes by behavioral change, reverting to pre-epidemic behavior will lead to a rapid rise in cases – the case in most urban and suburban counties. We find the cell phone data likely provides unreliable measures in many rural counties. We also find that behavior is unable to explain the epidemic in some counties, however these counties appear to be near highly populated counties with large numbers of cases, suggesting substantial county to county spillover. This suggest county-by-county modeling is likely insufficient for some locations. Finally, isolating infectious individuals is of first order importance. However, allowing recovered individuals, who are assumed immune in the model, to return to a baseline schedules has modest effects on the epidemiological dynamics in most counties, but a strong case reduction effect in a few places. This has implications for how scarce PCR and serological tests are deployed to help plan for reopening the United States.

## Analysis

### *R*_0_ *Analysis*

*R*_0_ is the number of the cases the index case would cause in a naїve population. It is measured at the moment an epidemic starts and is a function of behavior. So, *R*_0_ varies as baseline time use varies. After fixing *β*, the biological transmissibility parameter (see methods), we plot the variation in the *R*_0_ that would occur as a function of time allocated to public prior to the epidemic (mean time from January 1 through February 15)(Fig 1). This illustrates the importance of the behavior in *R*_0_ estimates. The two counties with the lowest *R*_0_ are suburban, Forsyth County, Georgia (*R*_0_ = 2.53) and Rockwall County, Texas (*R*_0_ = 2.62), followed by the rural Arthur County, Nebraska (*R*_0_ = 2.70). However, the right tail is inflated by rural counties where people may be spending more time alone (e.g., driving), but away from their homes and that have low device counts (see methods) and likely poor cell phone coverage. Historical driving time patterns are not useful to check this assumption because of congestion in urban areas during non-epidemic times. The right tail is dominated by counties like Loving County, Texas, Lake and Peninsula Borough, Alaska, and North Slope Borough Alaska. Many of these places are not reporting cases. The distribution in Figure 1, which has a mean *R*_0_ = 4.3 and median *R*_0_ = 4.2 aligns with other estimates of *R*_0_ for the United States, e.g., a national mean of 4.0, which was calculated early in the epidemic after some behavioral adjustment (Gunzler and Sehgal, 2020). The match is especially good considering that the right tail in Figure 1 is likely inflated.

**Figure 1.**
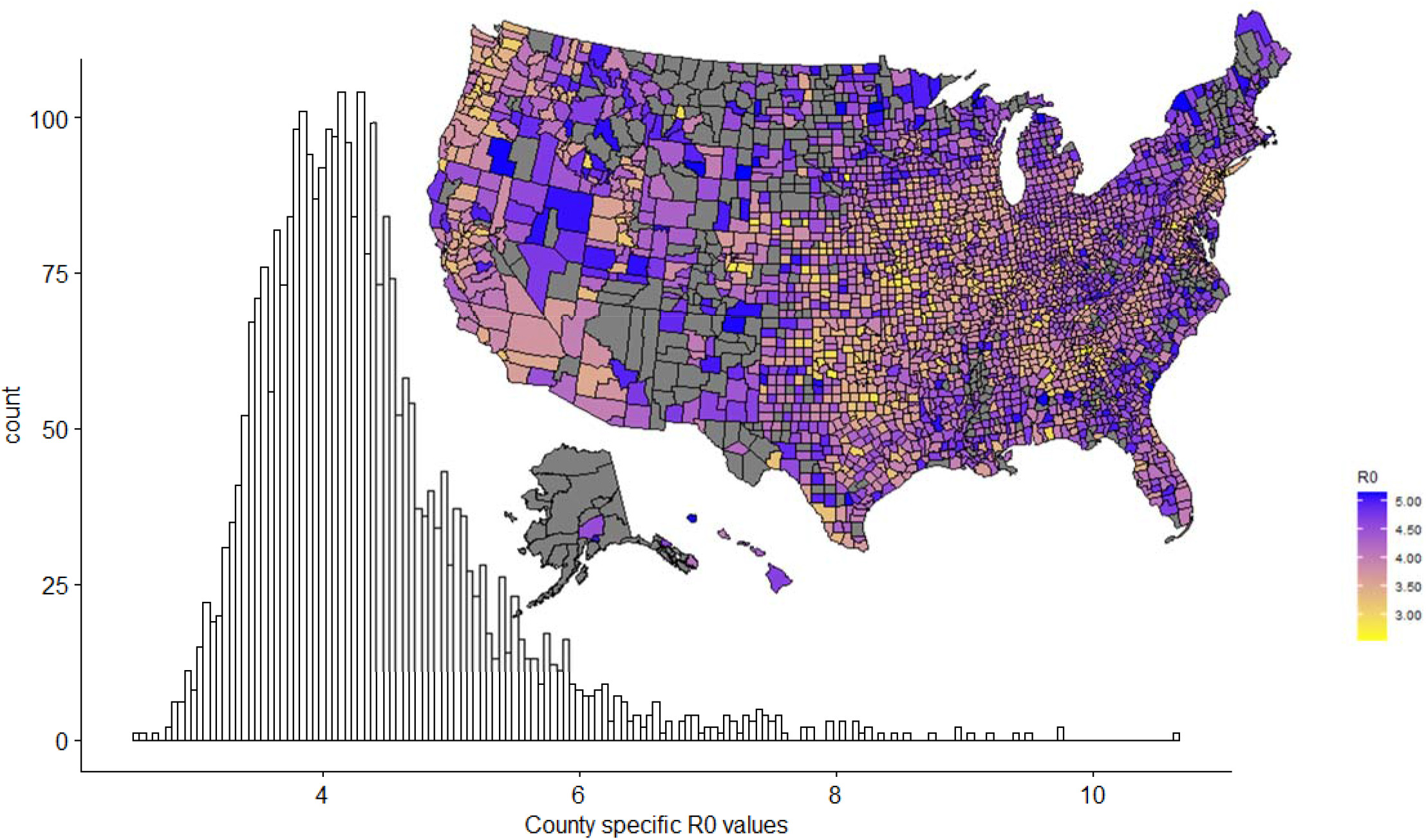
Distribution of expected *R*_0_ values for the United States. Different assumptions of the average *R*_0_ are horizontal shifts in the distribution. Grey counties are returning *R*_0_ > 5.2, this likely to do high driving times in remote areas or small cell phone sample size and are believed to be unreliable. However, *R*_0_ values for rural counties in general are likely unreliable – this is a caution for using cell phone data.

The results seem most credible for suburban, followed by urban counties. Outside of rural counties, *R*_0_ generally increases with density and urbanization. The local *R*_0_ can be quite variable. For example, the five boroughs of New York City are New York County (Manhattan) *R*_0_ = 5.1, Bronx County *R*_0_ = 4.0, Kings County (Brooklyn) *R*_0_ = 3.9, Queens County *R*_0_ = 3.5, and Richmond County (Staten Island) *R*_0_ = 2.9. Our results for rural counties based on cell phone data should be viewed with a high level of caution as being out of the house might not equate to mixing and transmission. Cell phone tracking data is likely less useful for understanding the epidemic in very low population density counties, at least without a substantial recalibration because how time spent out of the house translates into transmission contacts is likely substantially different.

### Epidemic Trajectories

Many, but not all, counties are “bending the curve”, as defined by having few cases over the last five days conditional on revealed behavior relative a simulation based on average baseline behavior, see methods (Fig 2, Fig 3). However, there are some counties where cell phone data suggests that behavior has hardly changed, e.g., New York County, New York (Fig 2 panel A) and Rockwall, Texas (Fig 2, panel E). Indeed, Rockwall, Texas suggests that the baseline behavior may be too conservative if the correct counterfactual scenario is that people would have increased time outside the home because of seasonal changes. Other counties are bending the curve enough to have experienced a peak in cases conditional on current behavior e.g., Queens County New York (Figure 2 panel B) and Forsyth County, Georgia (Figure 2 panel F). These counties have “flattened the curve,” as measured by declining active infectious individuals over the last five days. Some counties can have flattened curves or are past the epidemiological peak, but without behavioral change that flattened the curve, e.g., Custer County, Nebraska and Union County, New Mexico (Fig 3). We note, however, many of these counties have not actually experience epidemics, and the results stem from assuming an epidemic starts in every county once COVID-19 is reported in the state (see sensitivity analysis). There is a third category of county, where behavior is reducing the growth of infectious individuals, but infections are still growing, e.g., New Haven County, Connecticut (Figure 2 panel C) and Williamson County, Texas (Figure 2 panel D).These counties have “bent the curve,” but not flattened it. In all counties it is too early to resume “business as usual.” Even the counties with low *R*_0_ values, e.g., Forsyth County, Georgia need to maintain distancing for at least a month (Figure 2, panel F). Moreover, Forsyth County, Georgia illustrates the danger of stopping distancing too early (yellow curve). An important caveat is that the model does not account for interconnections among counties. Therefore, while Queens County and Forsyth County are on a trajectory for a near end of the epidemic, if behavior reverts to baseline and new infectious individuals are introduced into the county, then the epidemic will start again. This is why our model suggests some tightly connected counties, e.g., Queens County, are on a better trajectory than experience suggests.

**Figure 2.**
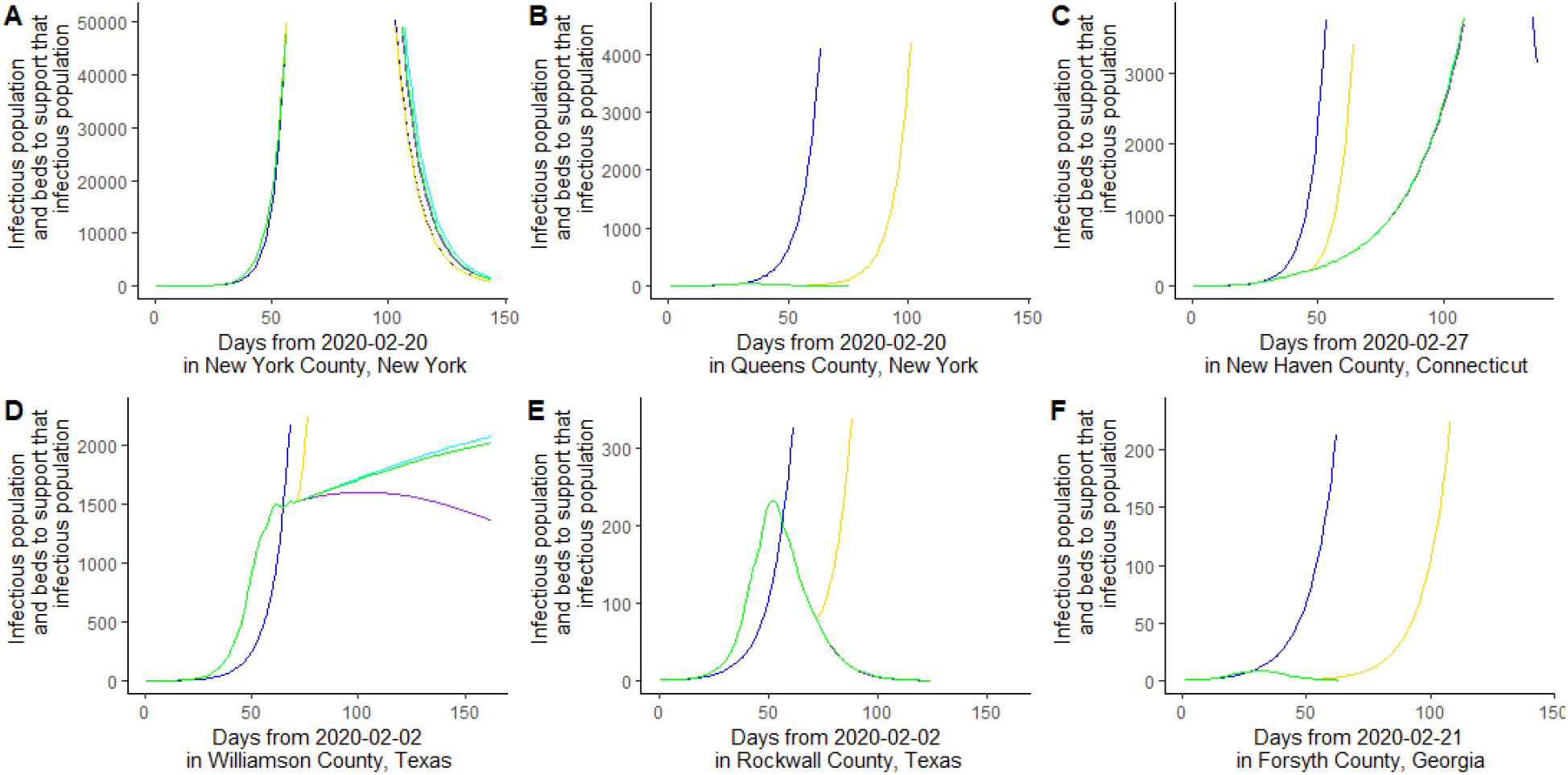
Illustrating three types of epidemiological trajectory, blue continued baseline behavior, yellow is based on cell phone data then reverting to baseline, green is continuing the current behavioral pattern, purple dots show what happens if recovered are allowed to revert to baseline behavior, and cyan is if both recovered and susceptible revert to baseline behavior. Panel A is New York County, New York where if anything people are spending more time in public. Panel B is Queens County, New York, which has already created a hump. Panel C is New Haven County, Connecticut, where behavioral shifts are reducing spread but not “flattening the curve.” Panel D is Williamson County, Texas. Panel E is Rockwall County, Texas. Panel F is Forsyth County, Georgia.

**Figure 3.**
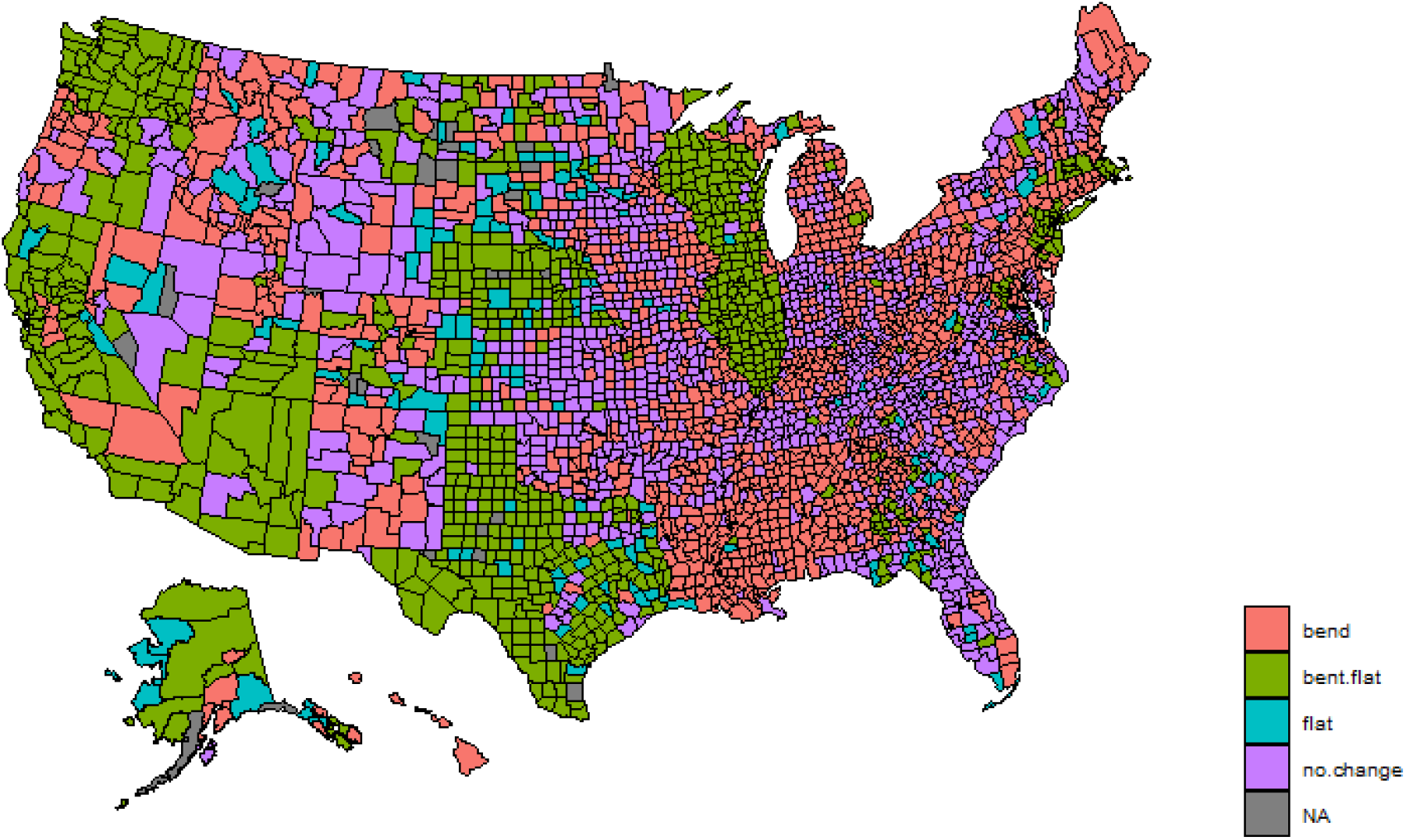
Map of the status of different counties with respect to the behavioral effects on the COVID-19 epidemic, bend means that behavioral shifts are reducing cases, but cases are still growing; bent.flat means that the behavioral shift is great enough such that new infections are declining; flat means new infections are declining but not because of behavioral shifts, no.change means that there is not a behavioral shift sufficient to reduce cases.

Overall there are 1,912 counties (61.5%) accounting for 74.5% of the US population that have bent the curve (Fig 3). There are 907 counties (29.5%) accounting for 30.8% of the US population that appear over the epidemiological hump. There are 755 counties (24.3%) accounting for 29.7% of the US population where behavior has flattened the curve. There are 1,036 counties where behavior has not shifted and may have exacerbated the epidemic. For the remaining of counties the differential equation solver does not converge because of changes in behavior and small population sizes.

### The effect of immunity and testing priorities

Testing is critical. There are two kinds of tests, PCR tests that can detect infectious individuals early for isolation and serological tests that determine who has antibodies. A subset of the serological tests coming online is aims to determine who likely has immunity. We focus on this later type of serological test. Our model helps identify where which kind of test should be prioritized.

PCR tests are a priority in areas where the curve has not bent or has not bent substantially, e.g., Bannock County, Idaho and New York County, New York. In these places, it is critical to begin identifying and isolating infectious individuals in order to more rapidly bring the COVID-19 epidemic under control. It is likely also important to maintain PCR testing in counties where the curve has not flattened.

Serological tests capable of identifying recovered and immune individuals (which are not yet available) are important, and the greatest benefits are in counties where getting recovered individuals back to baseline schedules reduces the greatest share of cases (Figure 6) coupled with those counties likely to experience the greatest hardships from infection (Maher et al., 2020). Our analysis suggests where the greatest percent and share of cases are avoided is correlated. The reason is that these counties are most likely to be bending, but not flattening the curve, and where there is a substantial number of recovered individuals that can make a difference, e.g., Williamson County, Texas (Figure 2, panel D). The model also shows that allowing susceptible and recovered individuals to baseline behavior (cyan curves in Figure 2) can lead to slightly more cases than maintaining recent behavioral shifts (green curves in Figure 2), in some cases, e.g., Williamson County, Texas. However, it leads to substantially few cases than returning the entire population to baseline (yellow curves in Figure 2). This suggests relatively low risk of a track, trace, and isolate program.

### Model Sensitivity

We test the sensitivity of the results to the infection recovery rate and the simulation start date. We reduce the recovery rate from 7.4 to 5 days. This reduces the national average *R*_0_ to 2.94 and median to 2.83 but does not change the shape of the distribution. This parameterization produces far fewer cases than observed in most counties. The patterns across the United States remains similar (Fig 5) to our primary calibration. However, more counties are near the peak of the epidemic, and therefore there is greater value in returning those with acquired immunity to baseline schedules. We find that 1703 counties (54.9%) are bending their curves accounting for 70.7% of the population. 743 counties have flattened curves (23.9% of counties and 34.9% of the population) with 579 counties behaviorally flattening the curve (18.6% of counties and 33.4% of population).

**Figure 4.**
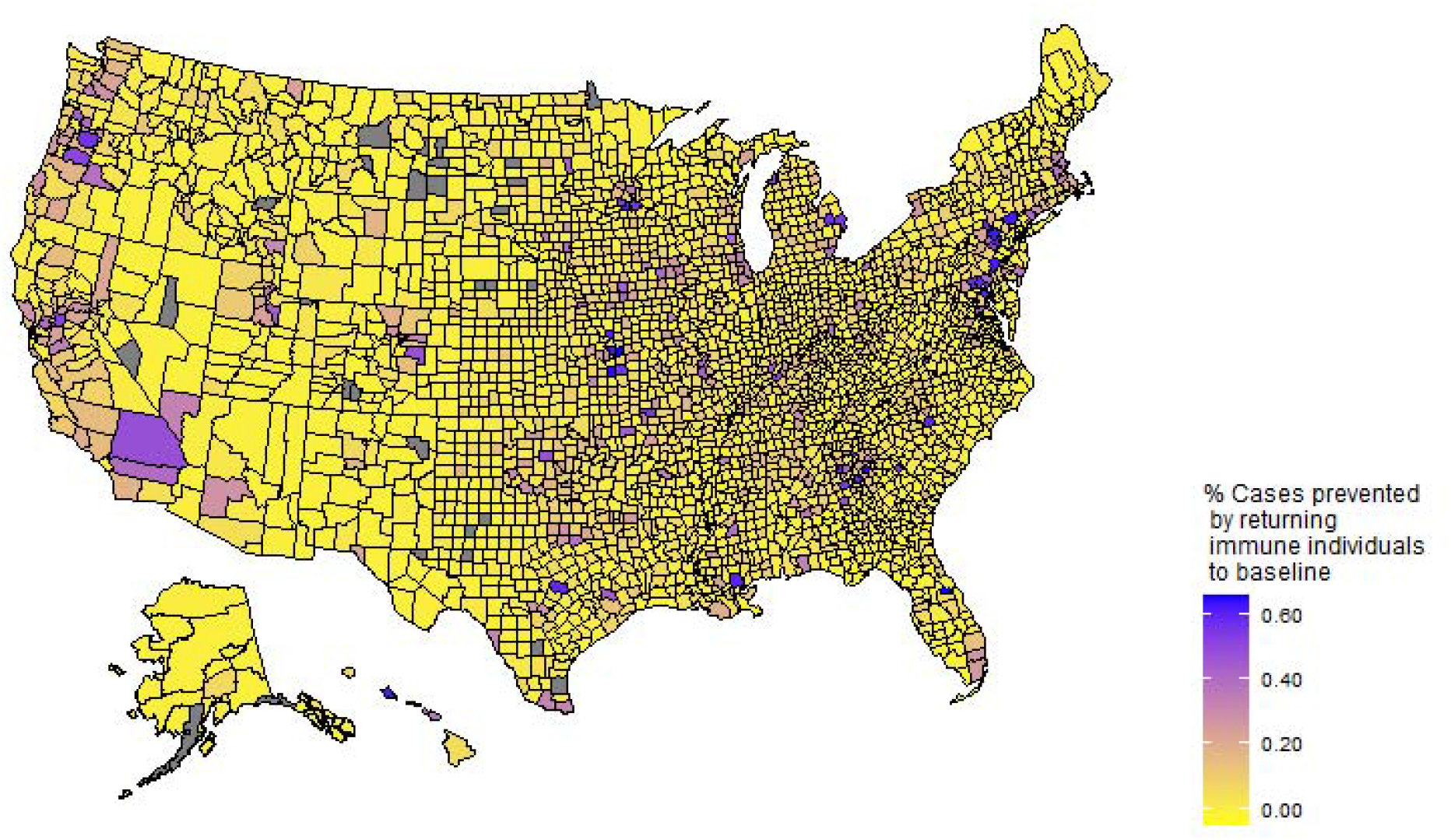
Showing the cases prevented if those with acquired immunity can return to baseline behavior

**Figure 5.**
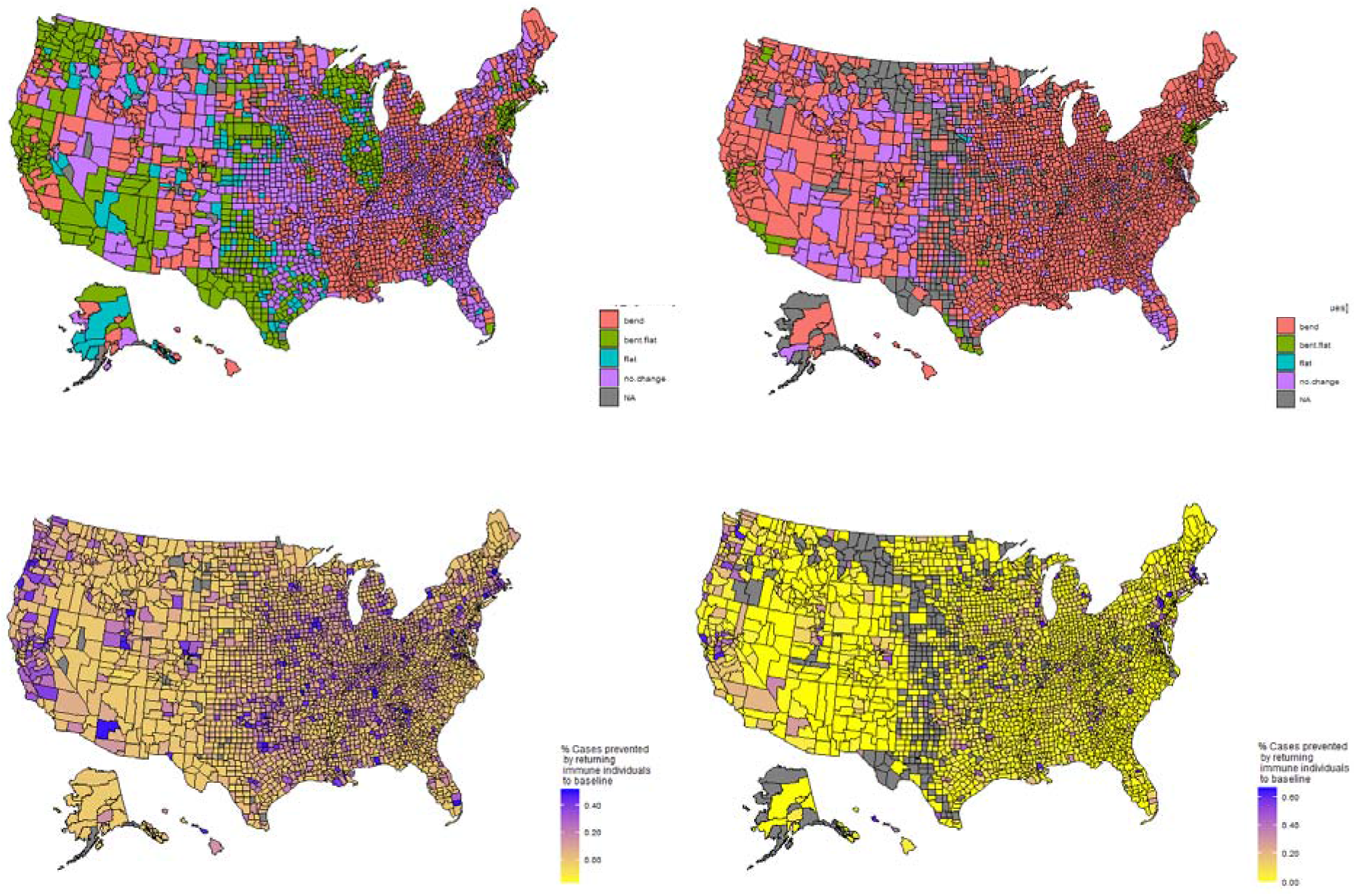
Sensitivity analysis for a recovery rate of 5 days, which reduces *R*_0_ (left column) and a started date 10 days before the county first reported (right column).

Second, we start every county’s epidemic 10 days before the first report in each county. For counties not reporting epidemics we use the date that the last county in the state first reported a first case. This leads to many counties where the epidemic does not happen because of changes in behavior that appear to respond to state wide or national case data. We find that 2382 counties (86.1%) are bending their curves accounting for 91.2% of the population. 81 counties have flattened curves (2.9% of counties and 20.6% of the population) with 78 counties behaviorally flattening the curve (2.8% of counties and 20.6% of population). This illustrates the simulations results depend heavily on the assumed start of the epidemic. Most counties are simpler earlier in their epidemics under this assumption, but this requires assuming a more disconnected country. Flattening the curve appears to require some saturation of susceptible individuals and sufficient time. Nevertheless, the overall pattern is robust, particularly with respect to the where returning recovered individuals to baseline behavior helps reduce cases.

## Discussion

The ability to couple near real time behavioral data with demographic data to understand how behavior is shaping the COVID-19 epidemic is critical for analyzing public health measures prioritizing testing, and developing back-to-work plans. By adapting the common SEIR framework to a dynamic time use structure that enables differential behavior by health we characterize the heterogeneity of COVID-19 epidemic across the United States. The model is sensitive to initial conditions and parameterization, but still produces robust patterns. Furthermore, the more the model is easily adjusted for local planning, and the analysis still provides an overview of the current situation.

A substantial share of the behavior in US counties is sufficient to have flattened the curve (Fig 3) or is on a trajectory do so. Flatten is a function of how long the epidemic has been going, behavior, the share of single person households, and population. Indeed, some counties experience flatten curves without behavioral shifts. Not all counties modeled as flattening the curve are actually experiencing flat curves. For example, Fairfield County Connecticut has experienced far more cases than the model predicts, but this county is tightly connected to New York County, New York, which has experienced a largely number of COVID-19 cases. This shows how large well-connected counties can swamp the local behavior of surrounding areas. It will be important to account for these connections when considering back to work planning. Nevertheless, a consistent pattern for most counties is that reverting to baseline behavioral will lead to a rapid rise in cases.

There are a number of remote rural counties where if the epidemic took hold at the same time as the rest of their states, then irrespective of behavior the cases may already be on a decline. However, the model is likely less reliable for rural counties because the measure of time in public is likely not equivalent to that in suburban and rural counties. Time in high risk locations, maybe a better measure than time at home. These are more likely to be complements (in the probabilistic sense) in suburban and rural areas, but less so in rural areas. Results from rural counties should be treated with extreme caution.

For counties still experiencing increasing numbers of infectious individuals (red and purple in Fig 3), it is important to enhance the isolation of infectious individuals to reduce new cases. PCR testing should be prioritized in these counties. Moreover, distancing measures will likely have to be maintained longer in these counties. Indeed, counties that are bending the curve, but not flattening it will experience the longest epidemics. These will be the most difficult counties to get back to work, but these counties will likely experience less mortality (perhaps because of the arrival of therapeutic medicines) than if they reverted to baseline behavior.

Getting America back to work is important to reduce economic hardships but must be approached with caution to prevent excess mortality. There are hopes that serological testing will enable those with acquired immunity to return to work. However, once a credible serological test is available, it will likely be in short supply. Maher et al. (2020) provide information on which counties are likely to suffer the greatest hardships conditional on COVID-19 epidemics. We show that an added benefit of disproportionally increasing the share of those with acquired immunity in public is that it also helps reduce cases – a double benefit. Reducing the cases in large connected counties likely has spillover benefits to surrounding and connected counties. These counties should be prioritized for serological tests capable of identifying immunity, once such tests become available.

We have focused on the first-order concern of targeting by health status. Models that make use of time-use data can be further refined by including other risk factors such as age, household size, and occupation. Such models are under construction. Nevertheless, two important caveats to smartphone driven time use models are that they likely perform best is suburban areas, then urban areas and worst in rural areas. Furthermore, time-use data for children is very important but exceedingly hard to gather. This should be considered as the evidence mounts that schools for young children may be one of the safest locations to open first (Viner et al., 2020).

## Methods: Model structure and calibration

We modify the standard SEIR (susceptible, exposed, infectious, and recovered) compartmental modelling structure in two ways to model social distancing. First, the model needs to account for in-home transmission, we do so by simplifying the age-and-household-structured model of (Bayham and Fenichel, 2016). Second, we modify the contact structure to allow for time varying time allocation and conditional proportional mixing (Fenichel, 2013; Fenichel et al., 2011). Additionally, we modify the disease induced mortality so that it is a function of prevalence or active cases in order to reflect exceedance of hospital capacity.

The general model structure for population *N*(*t*) = *S*(*t*) + *E*(*t*) + *I*(*t*) + *R*(*t*) at time *t* is

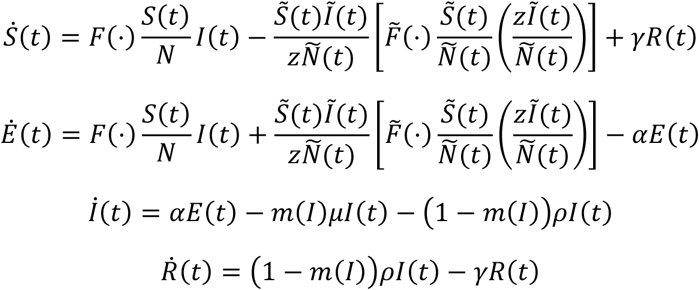

We suppress time when doing so does not cause confusion. The specification of other parameters primarily come from Wu et al. (2020) and are explained in Table 1, but we conduct sensitivity analysis. The function *m*(*I*) in an index function adjusts the mortality fraction as a function when the number of active cases exceeds the hospital capacity.

**Table 1.** Baseline national parameter values

| Parameter | Symbol | Calibrated Value | Source |
| --- | --- | --- | --- |
| 1/ Latency period | $\alpha$ | $5.0^{-1}$ | Wu et al. |
| 1/ Infectious period for those who recover | $\rho$ | $7.4^{-1}$ | Wu et al. |
| 1/ Infectious period for those who die | $\mu$ | $4.0^{-1}$ | Wu et al. |
| 1/ Period of acquired immunity | $\gamma$ | $9.13 \times 10^{-4}$<br>(3 years) | |
| Transmission parameter | $\beta$ | $1.33 \times 10^{-3}$ | Bilinski et al. (2020) & Safe Graph data |
| Mortality share when hospital capacity is not exceeded | $m(I)\vert I < capacity$ | 0.004 | https://ourworldindata.org/coronavirus#what-do-we-know-about-the-risk-of-dying-from-covid-19 |
| Mortality share when hospital capacity is exceeded | $m(I)\vert I < capacity$ | 0.04 | https://ourworldindata.org/coronavirus#what-do-we-know-about-the-risk-of-dying-from-covid-19 |

The first transmission term, 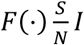, models transmission outside the home. It is the standard transmission function where 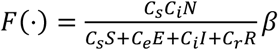 is the conditional proportional mixing transmission function. Lower case letters index their associated epidemiological compartments, and *C*_*j*_ is the amount of time an individual case *j* spends outside the home. In this formulation *β* is the probability that a susceptible individual becomes infected per minute contact with an infectious individual.

When people avoid public spaces, they reallocate time to home. The second transmission term 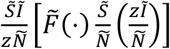 models infections within the home. *Ñ*(*t*) = (1 − *σ*)*N*(*t*) where *σ* is the share of the population that lives in single person households. Other epidemiological state variables are defined similarly. Transmission cannot occur in a single person household. The term in the square brackets is transmission within the representative household. 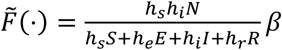, where *h*_*j*_ is the amount of time that an individual in health class *j* spends in the home interacting with others in the household. We assume there is zero marginal transmission probability during sleep and remove a six hour sleep duration. 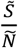 is the density of susceptible individuals in the representative household. 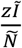 is the expected number of infectious individuals in houses of the representative size *Z* after eliminating single person households. 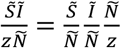 is the expected number of households that have infectious and susceptible individuals. Assuming infectious status is independent of household size, the product of the first two factors on the right-hand side is the joint probability of a house having a susceptible and an infectious individual. This likely overestimates household transmission, which means that measures of distancing effectiveness are a lower bound. The third factor is the number of households. This makes home relatively safe when prevalence, (*Ĩ/Ñ*), is low.

Ultimately, it is easier to think of *C*_*j*_ = *C*_0_ − *C*(*t*) and *h*_*j*_ = *h*_0_ + *C*(*t*), where *C*_0_ and *h*_0_ are the pre-epidemic allocation to time outside and inside the home, with *C*(*t*) the shift from outside the home to inside the home.

We do not attempt to provide realistic models of mortality. Without an age structure it is unreasonable to accurately model deaths in a representative agent model when deaths are heavily skewed toward the elderly. This is left for future work.

### Calibrating Transmission Parameters

*R*_0_ is often called a fundamental epidemiological parameter (Chowell and Brauer, 2009; Cohen and Enserink, 2009), but it is well known that, *R*_0_ confounds the biological features of pathogen with human behaviors that lead to transmission (Auld, 2003; Fenichel et al., 2011; Mossong et al., 2017; Springborn et al., 2015; Towers and Chowell, 2012; Zagheni et al., 2008). Therefore, *β* is the more fundamental parameter that we expect to be approximately constant irrespective of local behavior. The derivation of *R*_0_ involves taking the limit as *I* → 0 (Chowell and Brauer, 2009). The household transmission term goes to zero much faster than the public transmission term and has limited impact on the calculation of *R*_0_. Therefore, *β* = *R*_0_*ρ*/*C*_0_.

Bayham et al. (2015) calibrate time use with the American Time Use Survey (ATUS) (Hofferth et al., 2020). Using the ATUS data we find that the average American spent 351 minutes outside the home per day in the years prior to the COVID-19 epidemic, i.e., *C*_0_ = 351, with *h*_0_ as the residual of the 1,440 minutes in a day (counting time spent in one’s private car as time in the home), Figure (S1). However, the ATUS data are not updated regularly enough to be useful in the rapidly evolving COVID-19 epidemic.

Safe Graph provides near real time (three day lag) estimates of time spent in home based on anonymized cell phone location data (Safe Graph, 2020, www.safegraph.com). These data provide the median time that cell phones are recorded in the phone’s home location aggregated to census block group (600 to 3,000 people). The data appear more reliable when aggregated to the county level. For the average county, Safe Graph tracks an average of 5,639 devices (Fig S2). According to the Safe Graph data, the average American (population weight by county average) spends 669.7 minutes at home per day. To calculate time in public, first we remove six hours of time as sleep that is unlikely to lead to new contacts (Bayham et al., 2015). This leaves 410 minutes per day in public, which is similar to the ATUS data after accounting for the 65 minutes the average American spends in a private car per day (Fig 1). We use Bilinski et al.’s (2020) mean estimate of *C*_0_*β* (which they call *vp*) for Alameda, Contra Costa, Marin, San Francisco, San Mateo, and Santa Clara Counties in California, and divide by the population weighted *C*_0_ for those counties of 372.0 minutes to determine *β* = 1.33 × 10^−3^*β* is interpreted as the probability of infection per minute contact with an infectious individual.

County level hospital capacity is available from Homeland Infrastructure Foundation-Level Data (https://hifld-geoplatform.opendata.arcgis.com/. In our analysis we assume that 20% of cases will require hospital care. It is likely the far fewer cases are actually hospitalize, however disease induced mortality is not affecting the qualitative results of the model.

### Simulation Scenario

We consider five epidemiological scenarios for every county in the United States. We assume the epidemic begins in every county 10 days prior to the first reported case in the state with 5 infectious individuals. First, we build a baseline scenario that is based on the average time use between January 1, 2020 and February 15, 2020. This does not account for seasonal changes in behavior. If people are more likely to spend time out of the home as the weather warms, then our baseline is underestimating the counterfactual epidemic. Second, we use the baseline behavior until the first case is reported in the state, and then impose the time series of actual time use in each county, but allowing individuals to revert to baseline behavior when the data end (currently April 13^th^). By imposing the actual behavior we account for any public policy and compliance with that policy. Third, we take the mean of the last five days of time use and continue that behavior. Fourth, we mimic the third scenario but allow those recovered with immunity to return to baseline when the behavioral data end. Fifth, we allow susceptible individuals and recovered individuals to return to baseline behavior, which is equivalent to reducing the time in public of exposed and infectious individuals, though not complete isolation.

We classify counties into four groups: non-behaviorally responsive, curve benders, those with a flat curve, and curve flatteners. Curve benders are counties where the number of infectious individuals over the last five days of observed Safe Graph time use data is less than using the baseline counterfactual scenario, whereas for non-behaviorally responsive counties this condition does not hold. A county has a flat curve if the slope of the infectious individuals curve over time is less than or equal to zero over the last five days of Safe Graph time use data. It is possible that counties have a flat curve, but have not adapted behavior. This happens in small counties. Therefore, counties classified as curve flatteners satisfy the flat curve condition and the curve bender condition.

We compute the number of avoided cases over the remainder of the epidemic by allowing recovered individuals to revert to baseline behavior relative to the case were current behavior is maintained for all.

### Sensitivity Analysis

Reliably calibrating a model of COVID-19 is challenging given the large number share of undetected cases (Bendavid et al., 2020; Stock et al., 2020) and the detection rate varies through time. Furthermore, the United States data does not provide a reliable record for calibration. The distribution of reported cases 12 days after the first case in a county does follow a reasonable pattern (Fig S3). We perform two sensitivity analysis. First, we reduce the recovery rate from 7.4 to 5 days. Second, we start every counties epidemic 5 days before the first report in each county. For counties not reporting epidemics we use the date that last county in the state first reported a case.

## Data Availability

Data is either publicly or available for COVID-19 researchers from Safe Graph. Code will be made available following peer-reviewed publication (if not sooner).

---

**Figure S1.**
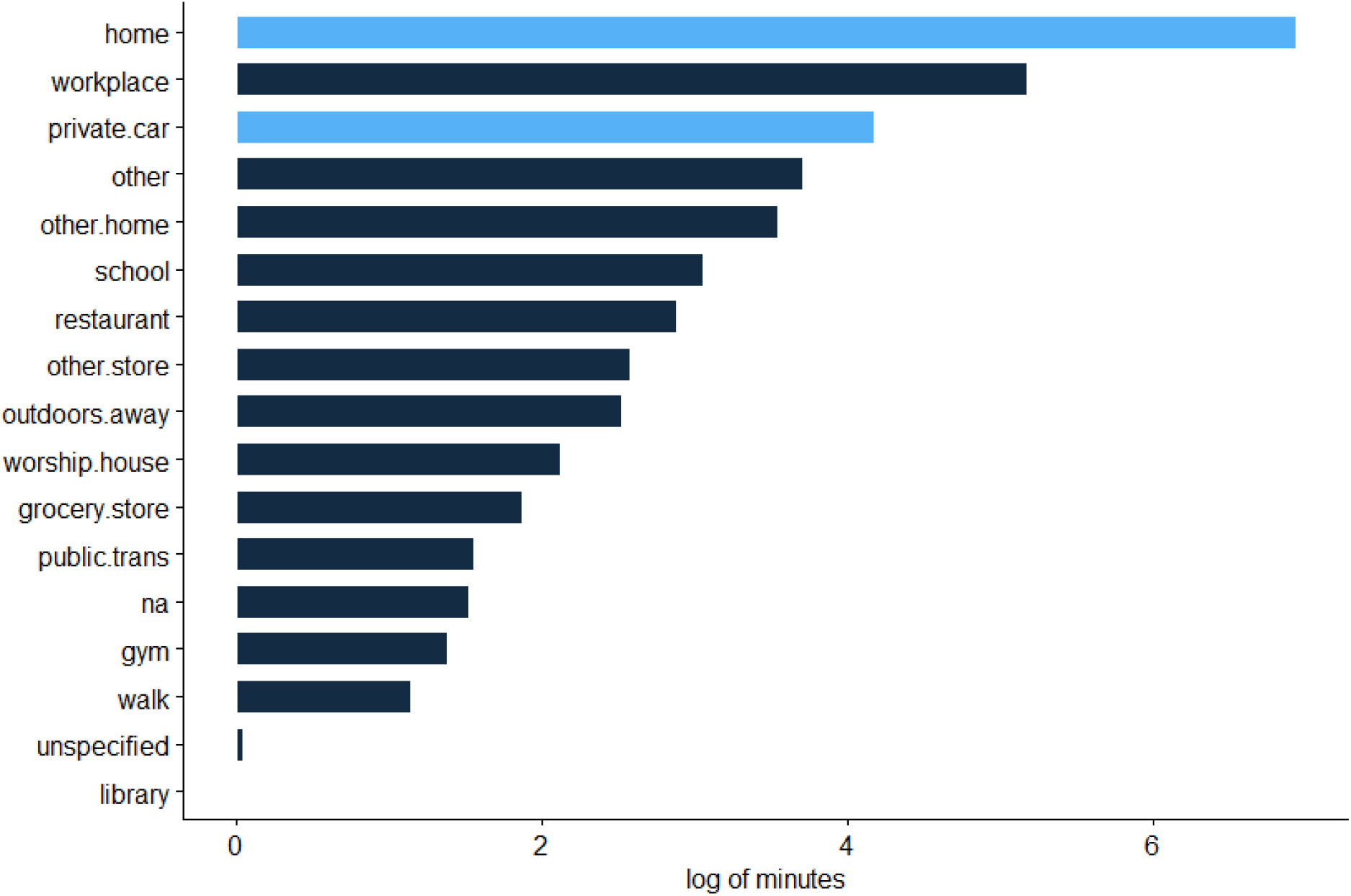
The distribution of time use by the representative American. Light blue is considered home time, and dark blue is considered time in public.

**Figure S2.**
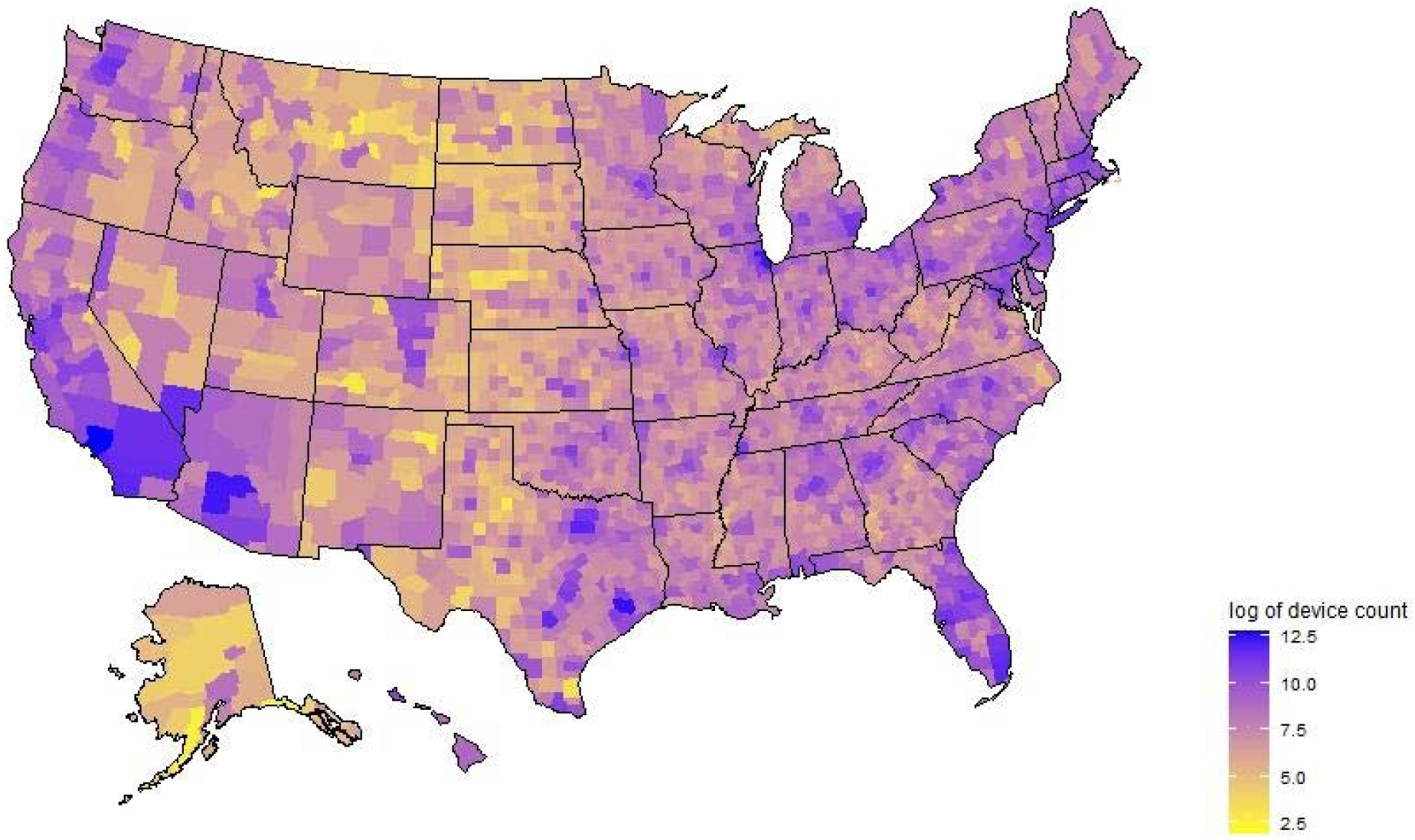
Safe Graph smartphone coverage.

**Figure S3.**
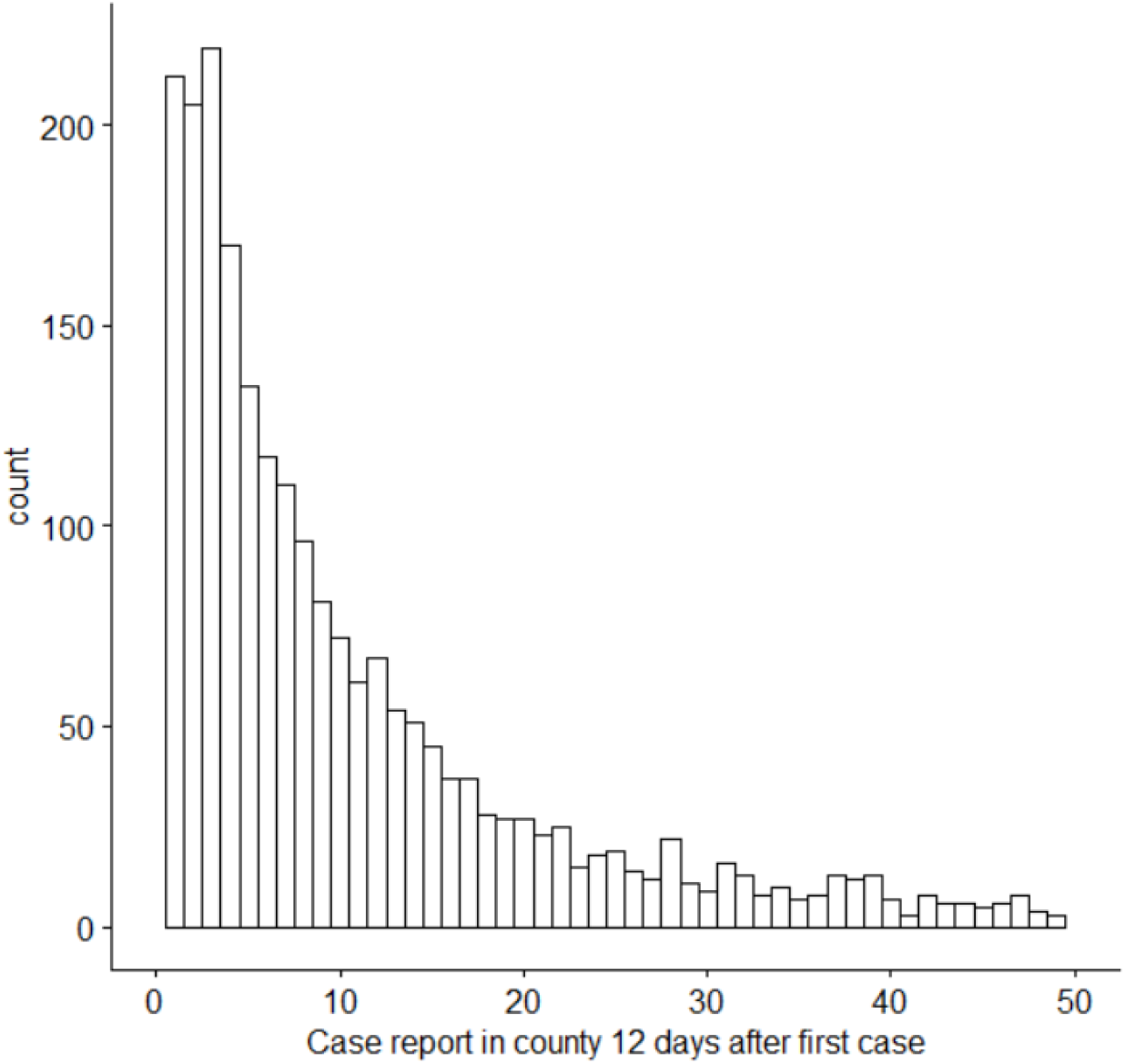
The distribution of cases reported in a county 12 days after the first case in that county is reported based on data from the New York Times. The right tail is truncated, but we would have expected a mode much greater than 2.

